# A novel computational approach to reconstruct SARS-CoV-2 infection dynamics through the inference of unsampled sources of infection

**DOI:** 10.1101/2021.01.04.21249233

**Authors:** Deshan Perera, Ben Perks, Michael Potemkin, Paul Gordon, John Gill, Guido van Marle, Quan Long

**Affiliations:** Department of Medicine, Cumming School of Medicine, University of Calgary and Alberta Health Services, Calgary, AB T2N 4N1, Canada; Department of Biochemistry & Molecular Biology, Alberta Children’s Hospital Research Institute, University of Calgary, Calgary, AB T2N 4N1, Canada; Department of Microbiology, Immunology, and Infectious Diseases, Cumming School of Medicine, University of Calgary, Calgary, AB T2N 4N1, Canada; Department of Medical Genetics, and Mathematics & Statistics, Alberta Children’s Hospital Research Institute, O’Brien Institute for Public Health, University of Calgary, Calgary, AB T2N 4N1, Canada

**Keywords:** COVID19, SARS-CoV-2, molecular phylogenetics, transmission inference, transmission network, infectious diseases

## Abstract

Infectious diseases such as the COVID19 pandemic cemented the importance of disease tracking. The role of asymptomatic, undiagnosed individuals in driving infection has become evident. Their unaccountability results in ineffective prevention. We developed a pipeline using genomic data to accurately predict a population’s transmission network complete with the inference of unsampled sources. The system utilises Bayesian phylogenetics to capture evolutionary and infection dynamics of SARS-CoV-2. It identified the effectiveness of preventive measures in Canada’s Atlantic bubble and mobile populations such as New York State. Its robustness extends to the prediction of cross-species disease transmission as we inferred SARS-CoV-2 transmission from humans to lions and tigers in New York City’s Bronx Zoo. The proposed method’s ability to generate such complete transmission networks, provides a more detailed insight into the transmission dynamics within a population. This potential frontline tool will be of direct help in “the battle to bend the curve”.

## 1. Introduction

Public health programs have been repeatedly tested over many years by sporadic events caused by the emergence of novel pathogens. Poorly focused public health interventions or indeed the lack of any action have led to uncontrolled epidemics and in some instances global pandemics [1,2]. In the 21^st^ century alone, the world has experienced everything from the outbreaks of the Ebola virus and Zika virus, to Influenza viruses and the present pandemic from SARS-CoV-2, the causative agent of COVID19[1,3,4].

Whole-genome viral sequencing, a new tool for genomic epidemiology, is playing an increasingly important role in the investigation of many infectious disease outbreaks [5]. The current COVID-19 pandemic reinforces the potential importance of these approaches. The primary goal of any epidemiologic investigation is the mitigation and termination of disease spread. Analysis at the nucleotide level resolution brought about by sequencing technologies is now being widely used in the characterisation of causative pathogens and the identification of their transmission patterns [2,6]. Some viral genomes, in particular the rapidly mutating RNA viruses, generate sufficient genetic diversity for their use in the inference of the pathogen’s transmission. Therefore, genomic epidemiology has the potential to be both a feasible and useful tool to infer epidemiological dynamics solely through the use of viral genomes collected, often for diagnosis, across the timeline of an epidemic [1].

The complete and accurate inference of infection transmission networks enables health care professionals to implement effective protocols to mitigate disease spread [1]. However, in practice, such an approach is hampered by the inability to completely sample an entire population [7]. Unsampled sources of infection play a significant role in infectious disease transmission leading to the rise of unsuspected clusters of infection. They also impede precise estimates of the burden of infection in a population [7,8].

Many of the challenges in the COVID19 pandemic relate to the inability to adequately track virus spread within a community [9]. In COVID19 some infected individuals may be asymptomatic or present with delayed symptoms many days after they have been infectious to others. This leads to the presence of the “silent” spreaders within a community [10]. These individuals may explain in part the unprecedented infection rates, atypical clusters as well as unaccounted cross-species transmissions seen in COVID-19 [8,11]. Therefore, a systematic way of predicting SARS-CoV-2 transmission networks is crucial to correctly estimate the number of asymptomatic or undiagnosed sources and ultimately reduce the number of those infected.

In this study, we evaluated a novel approach, using the rapid rate of viral mutation, with the ability both to infer transmission networks and to predict the presence of unsampled sources of infection. This high viral mutation rate, while challenging for vaccine development and viral identification, can be useful as a submicroscopic genomic map to help decipher the transmission of the pathogen from one host to the other [1].

A multitude of approaches aimed to infer inter-host viral transmission using these within-host evolutionary dynamics do currently exist. These include the work conducted by Didelot et al., Stapleton et al., and Xu et al. [7,12,13]. We have extended these well-established methods and our own pipeline [14] designed for estimating HIV transmission in a population, into a refined protocol aimed to infer the transmission of SARS-CoV-2. Through our work, we have been able to confirm key questions on the effectiveness of a country’s sampling practices, the success of social distancing, and how certain infected clusters are connected through the prediction of once unknown sources of infection. For instance, our approach sheds further light on the infection of the lions and tigers at the Bronx Zoo in New York City, USA whose source of infection raised much interest [15].

The strength of our proposed pipeline lies in its ability to infer the presence of unsampled sources of infection that may be contributing to infectious disease spread. This ability may assist in answering questions regarding the direction of transmission and how certain infected populations are connected. We believe that through a better understanding of the transmission dynamics, one would finally be able to get ahead of the curve.

## 2. Methods and Materials

### 2.1. Overview of the proposed pipeline

The proposed transmission pipeline, as shown in **Fig. 1** is composed of five steps: 1. Date extraction, 2. Multiple Sequence Alignment (MSA), 3. Parameterization and phylogenetic inference 4. Transmission tree generation and 5. Data visualization.

**Fig. 1.**
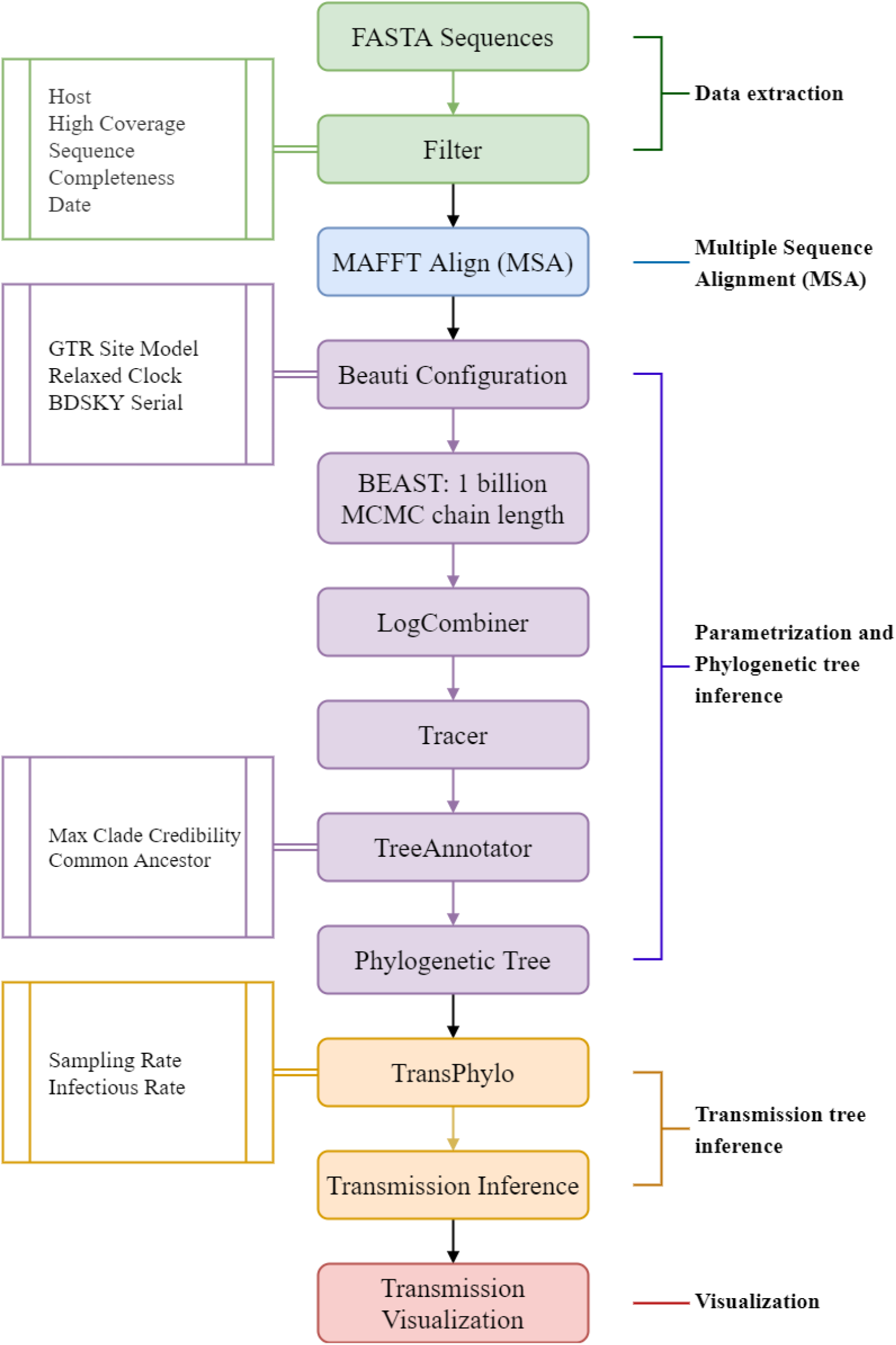
A detailed overview of the proposed five-step pipeline. The parameters that were optimised to fit the dynamics of the SARS-CoV-2 virus are depicted on the left.

Through multiple testing and iterations, this multistep process was optimized specifically for the SARS-CoV-2 virus. This enabled the generation of both sound transmission networks of COVID19 as well as the prediction of unsampled sources of infection.

### 2.2. The three scenarios of study

Our total analysis spanned across Canada, Germany, France, Africa, South Korea, Russia, and New York State and we focus on a detailed discussion of three main regions, namely Canada, Russia, and New York State. Each data set proposed a new set of challenges that had to be solved and, in each use case, our novel approach proved its versatility.

The Canadian dataset that we selected for analysis was concise. The sampling rate at the time of data collection which spanned from January of 2020 to July of 2020 was sufficient and the local governments had implemented approaches that appeared to control the infection. The selected dataset also spanned across multiple provinces. The dataset for the COVID19 pandemic in that time interval was well documented in Canada allowing us to easily validate our results. It represented a good real-world scenario to test our pipeline [16].

The second dataset collected in Russia from March of 2020 to September of 2020 was similar to that of Canada but with one significant change. The dataset from Russia appeared to be less sampled and therefore the system would have to predict a large number of unsampled sources. Simultaneously, it would have to correctly infer the directions of disease transmission connecting infected population clusters, based on the epidemiological data available in the GISAID database [17].

Finally, the New York State data also represented a series of obstacles that the pipeline would have to tackle. At the time of data collection from February 2020 to September 2020, the state was being considered a COVID19 epicenter had the second-highest cases of COVID19 in the USA by March 2020. It also had a large population of people traveling into the region and social distancing practices had not been strictly implemented [18,19]. It was also evident that much like the Russian dataset that New York State had a significantly less sampling rate in comparison to its rate of disease incidence [20]. Therefore, this would act as a potentially challenging data set for a pipeline like ours. To further test the robustness of our system we took measures to include the SARS-CoV-2 sequences obtained from the lions and tigers at the Bronx Zoo, New York. The sequences from the zookeepers were not present in GISAID and therefore were not included in the dataset to ensure the continuity of quality control standards of the data. We expected the pipeline to be able to infer the cross-species transmission in spite of this, enabling us to truly validate its efficacy.

### 2.3. Viral sequence collection and quality control

The selected data source was GISAID (https://www.gisaid.org/) [17]. Both FASTA sequence data and the corresponding epidemiological data were downloaded. It was ensured that the selected data was complete sequences with high sequence coverage. Using a bespoke filtering algorithm, sequences with complete collection dates and locations were selected ensuring compatibility for the generation of timed phylogenetic trees in the subsequent steps.

### 2.4. Multiple Sequence Alignment (MSA)

MSA was performed using Multiple Alignment using Fast Fourier Transform (MAFFT) [21]. MAFTT was the aligner of choice due to its robustness in handling large datasets as well as its speed and efficiency which comes with minimal cost to the accuracy of the alignment [22–24].

### 2.5. Phylogenetic parameter selection

Phylogenetic reconstruction was performed through Bayesian time-trees employing BEAST 2.6.2 [25]. The analysis was conducted with activated tip dates, a Generalized Time Reversible (GTR) site model with a gamma category count of 4, a relaxed clock model, and a Birth-Death Skyline Serial model as the prior. Parameter selection was influenced by our previous work on HIV transmission [14].

### 2.6. Phylogenetic tree generation

The total MCMC chain length stood at 1 billion which was performed as 10 separate runs each of chain length 10^7^. The individual runs were then merged using LogCombiner 2.6.2 and the validity of the MCMC run was evaluated using Tracer 1.7.1 by reference to the Estimates Sample Size (ESS) of each inferred parameter. It was ensured that the ESS was greater than 200 for each parameter. Subsequently, the phylogenetic tree was extracted through TreeAnnotator 2.6.3 using common ancestor node heights and a target tree type of maximum clade credibility.

### 2.7. Transmission tree inference

Transmission tree inference was conducted using the Bayesian program TransPhylo; a dedicated software designed to reconstruct transmission networks from timed phylogenetic data [7]. TransPhylo was particularly well suited for the scenario of COVID19. TransPhylo enabled the inference of transmission trees for an ongoing pandemic complete with unsampled sources of infection and the date of infection. The identification of unsampled sources was particularly important in identifying how certain clusters were connected and understanding the direction of disease transmission. TransPhylo was executed with parameters that represented generation times within 1 to 14 days with a median of 5.5 days and sampling times of 2 to 14 days with a median of 7 days. It should be noted that these parameters had to be varied, usually within these boundaries based on the region under study.

### 2.8. Visualization of transmission networks

Data visualization was conducted using Gephi 0.9.2 a specialized network analysis software [26]. Gephi’s built-in clustering algorithms namely Force Atlas 2 and Yifan Hu enabled us to identify population clusters in the transmission network as well as visualize the data in a comprehensible manner [27,28].

## 3. Results

Using the richness of the GISAID (https://www.gisaid.org/) [17] data, we tested and optimized our pipeline for the analysis of the COVID19 pandemic. Through consecutive tests, we have validated our pipeline in terms of its statistical soundness in addition to the endeavor of authenticating its predictions through rigorous fact-checking.

### 3.1. Statistical validation of Bayesian MCMC runs

The MCMC runs conducted in both the inference of the phylogenetic tree by BEAST and the transmission tree by TransPhylo were statistically validated by examining their trace diagrams. It was ensured that the BEAST generated results had Estimated Sample Sizes (ESS) above 200.

Certain parameter values were not entirely consistent with the available literature. Most of the analyzed regions had predicted reproductive numbers which ranged from approximately 0.2 to 0.3 while some of the larger data sets, Africa for example, had values between 0.4 and 0.8. Out of the ten different estimates of the reproductive number for each region, there would be outliers giving a much higher value, ranging anywhere from 1.0 to 2.5. When compared to literature, it was determined that the majority of these values were inaccurately small [29,30]. Through the analysis of 42 studies on three different databases by Billah et al., it has been determined the average worldwide reproductive rate ranged from 2.39 to 3.44 [29]. Regarding the specific reproductive rates in the examined regions, the following values were found from previously conducted studies: France - 2.72 to 6.32, Germany - 5.51 to 6.69, Russia - 1.30, China - 3.28, South Korea - 0.26 to 1.60, and the United States - 1.61 [29–31]. Regional reproduction values for African countries were unable to be determined due to the smaller data set encompassing a large geographical region. South Korea was the only region in which the predicted reproductive number was similar to previously published values [29]. Regarding the “become uninfectious rate” parameter, the predicted values were not close to those present in SARS-CoV-2 literature. The most likely reason for both these errors was due to the short time span of the sampled cases as well as the relatively small number of samples. Despite these results, it was also observed that down the line these errors had minimum to no effect when inferring transmission patterns and predicting infection dates.

In the event of TransPhylo, it was ensured that the trace diagrams indicated convergence by the end of the MCMC run. It was noted that based on the region of study the value at which convergence was reached varied.

### 3.2. Spread of COVID19 in Canada

The first reported case of COVID19 infection in Canada was seen in January of 2020 in Ontario, Toronto followed by reports of documented infections in several regions of British Columbia by early February followed by a massive uprise of cases in Quebec and finally to the rest of Canada by late March. During this time period, extensive sampling of infected individuals was carried out [32–34]. The transmission network diagram produced by our pipeline (**Fig. 2**) successfully predicted these transmission events.

**Fig. 2.**
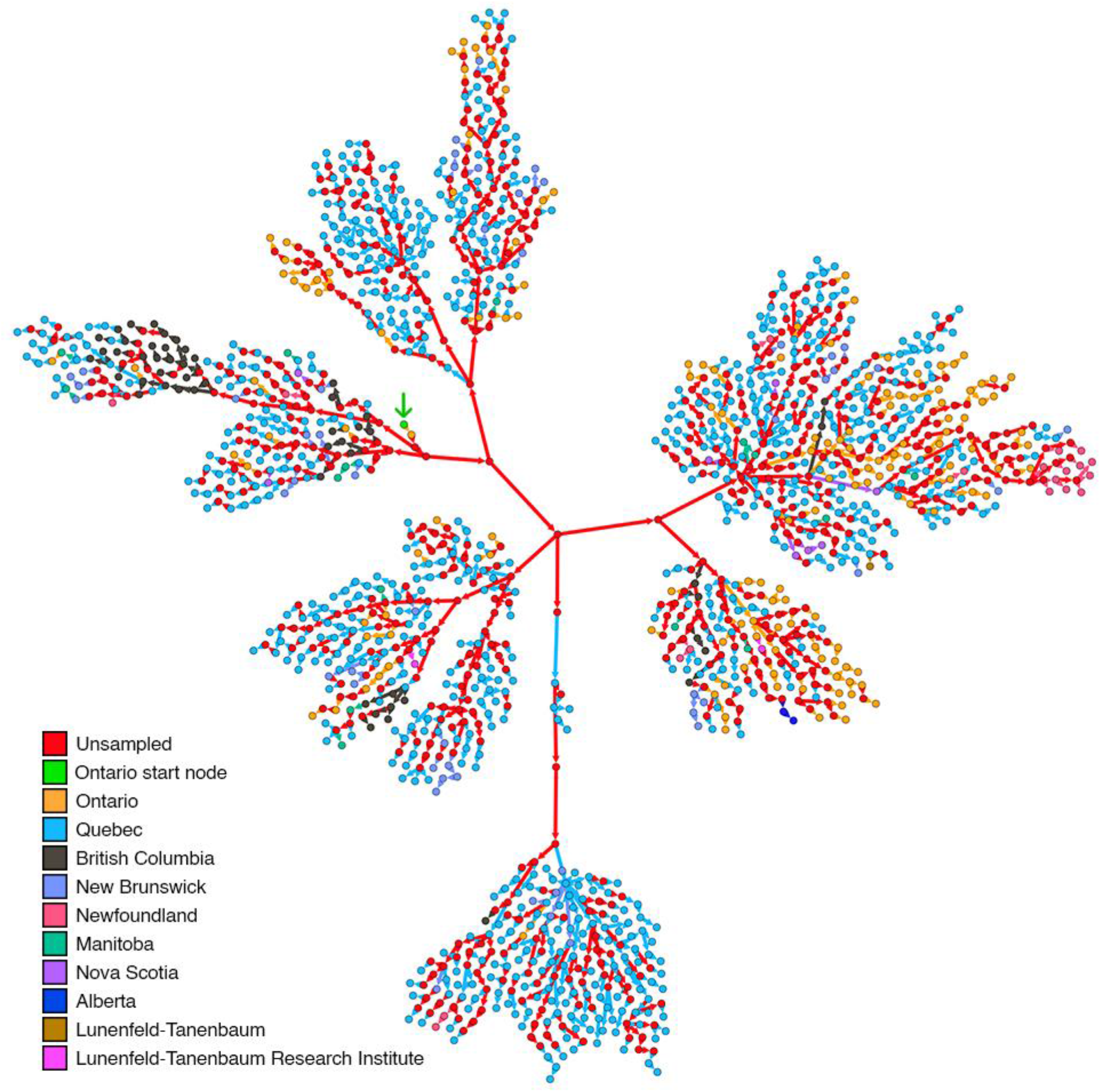
The transmission diagram obtained for the Canada dataset colour coded based on the region from which the sampled nodes were obtained. The first node of infection which has originated in Ontario has been marked by a green arrow, predicted unsampled sources of infection are coloured in red and the colour of the arrow depicts the recipient node. The diagram clearly depicts the effect of social distancing with clear isolation of the nodes of the same colour. The unsampled nodes (35.63% shown in red) help complete the transmission chain.

Due to the promotion of social distancing practices in Canada by mid-March the transmission diagrams show the clustering of nodes within the same region as regions started to slow the spread of COVID-19 infection [35]. This documents the success of the prevention protocols. This success is most evident when examining the transmission of COVID-19 in Newfoundland (which belongs to the “Atlantic Bubble” established on the 3^rd^ of July 2020)[36]. There were no in-province transmission events predicted for Newfoundland from March of 2020 to July 2020 suggesting the success of the Atlantic Bubble. Therefore, through the transmission diagram obtained from our pipeline, the potential effects of various infection control protocols can be easily visualized.

### 3.3. Spread of COVID19 in Russia

The transmission diagram of the Russian dataset (**Fig. 3**) also shares several similarities with the literature, despite the difficulty in making more concrete comparisons due to the lack of extensive epidemiological data. Kozlovskaya et al. [37] state that the first several cases identified in Russia were found in Moscow, and that national transmission may have started from there, and would have been bolstered by new arrivals to other cities. This is backed up by the presence of Moscow-derived samples appearing at a large portion of larger cluster junctions in our diagram, indicating travel that may have led to outbreaks in other cities such as Omsk and Saint Petersburg. Additionally, Komissarov et al. [38] have identified nine distinct transmission networks within the Russian Federation, making use of similar data obtained from GISAID [38]. Several of the transmission networks deduced by Komissarov et al. include transmissions from Moscow to Yakutia, from Krasnodar to Orenburg, and from Moscow to Sverdlovsk. These transmission pathways are compatible with the results obtained from our analysis, as TransPhylo did show transmission between the former two regions and transmission through four unsampled sources for the latter transmission pathway [38]. Additionally, several large outbreaks are seen in Saint Petersburg, since similar data has been used, these outbreaks may contribute to outbreaks at the Vreden Hospital which occurred somewhere from late-March to early-April [38].

**Fig. 3.**
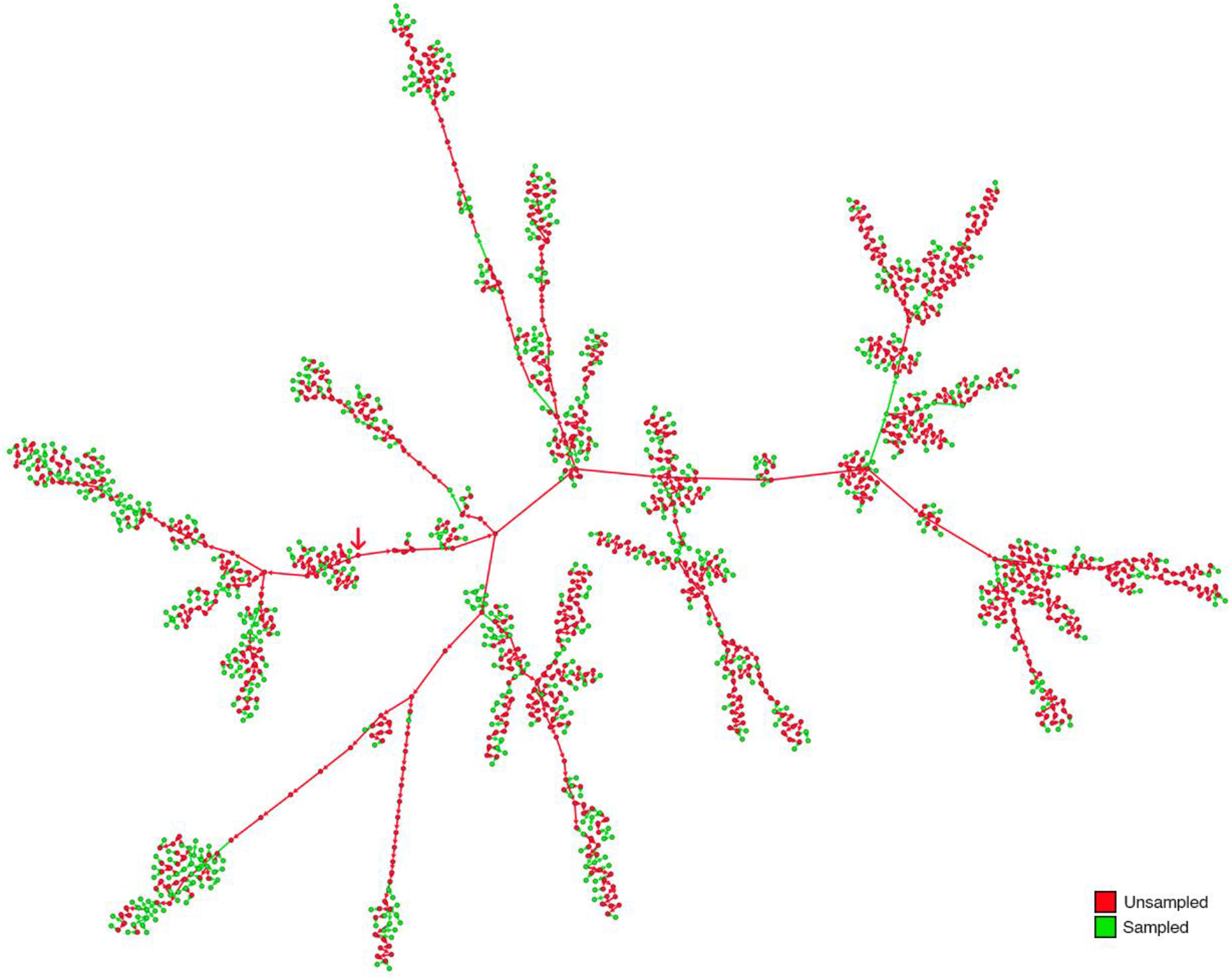
The transmission network obtained from the Russian data set has a considerably large number of unsampled sources (61.06% shown in red) in contrast to the sampled sources (38.94% shown in green). It was still able to accurately predict certain transmission events that were authenticated by the available literature. The source of infection is this network is a predicted unsampled node marked by a red arrow (center left)

### 3.4. Spread of COVID19 in New York State and tackling cross-species transmission

As stated, the New York State dataset was selected to further test the robustness of the proposed pipeline. The resultant transmission diagram (**Fig. 4**) appears to have an epicenter with a large number of unsampled nodes overtaking the number of sampled cases. The central node’s predicted infection date of December 2019 coincides with the world’s first reported cases of COVID19. Due to the large numbers of predicted unsampled sources of infection coupled with the lack of clustering of nodes from the same region, it can be concluded that on average, the individuals in New York State were more mobile and the seemingly available social distancing practices appeared not to limit the spread. The available literature on the spread of COVID19 in New York State seems to confirm these predictions [39–41].

**Fig. 4.**
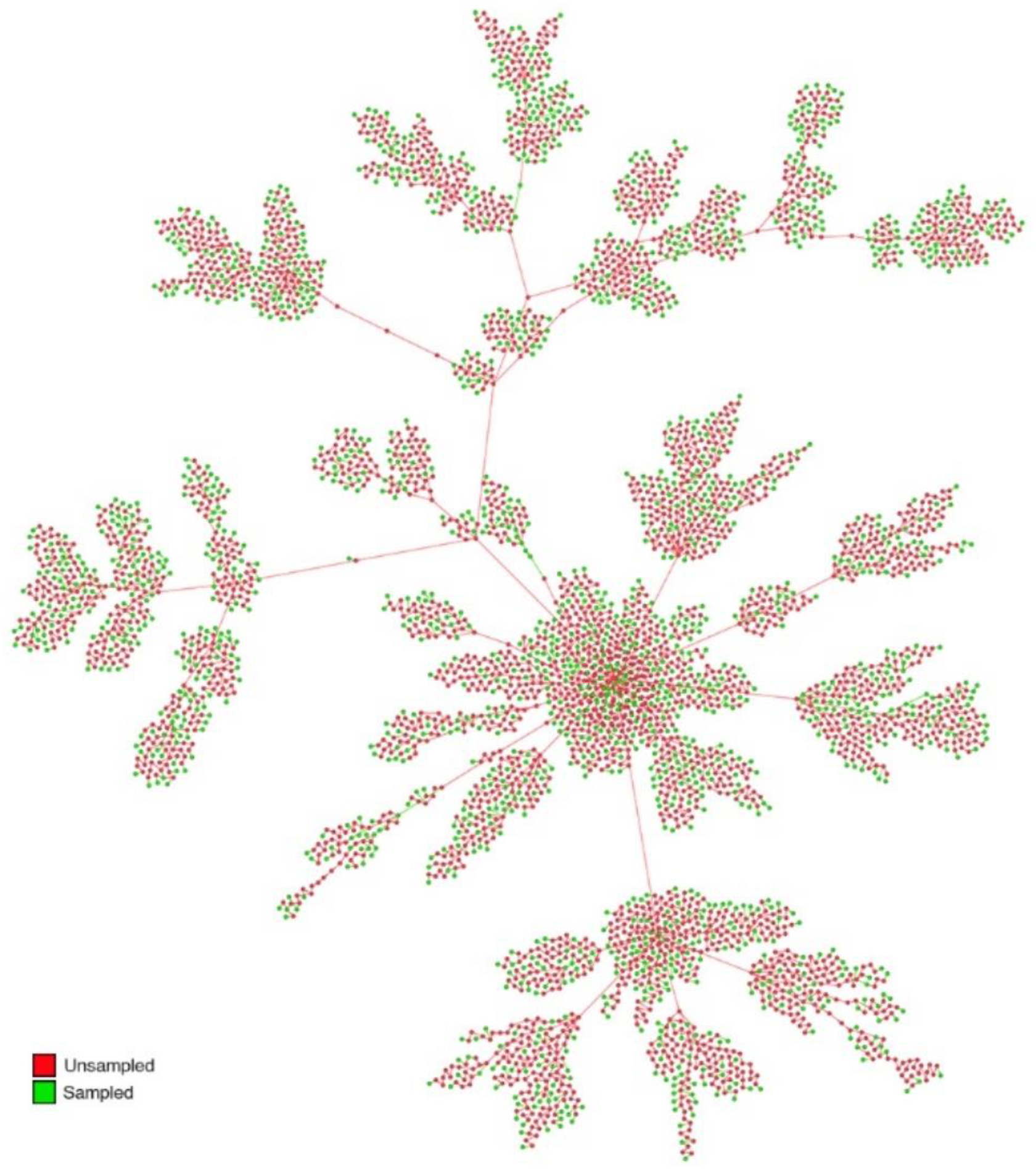
Transmission network diagram obtained from the New York State dataset. Here too the number of predicted unsampled sources (65.73% shown in red) greatly outnumber the number of sampled sources (34.27% shown in green) The clear formation of a central cluster of nodes can be seen due to a high number of introductions of the infection into the region from the outside.

In April of 2020 seven tigers and four lions at the Bronx Zoo, New York City were infected with SARS-CoV-2. The subsequent extensive clinical analysis showed that the transmissions were from a human to lion and human to tiger cross-species transmission [15]. Our pipeline was capable of making this inference solely from the genetic data (**Fig. 5a**). The pipeline was also able to accurately infer a series of unsampled sources of infection that arose from sampled sources and resulted in the infection of the lions at the Bronx Zoo. To confirm the validity accuracy of this prediction we analysed the epidemiological data of the nodes surrounding the lion’s and tiger’s data points (**Fig. 5b**). This revealed that these sampled sources were tested in laboratories all of which were less than 12 miles from the Bronx Zoo. These included the New York University Medical Center and QDX Pathology which are situated about 11 miles or a half an hour drive away from the Bronx Zoo. Assuming that all sources sampled at these sites lived in the area serviced by these laboratories, this information reinforces the accuracy and robustness of the pipeline.

**Fig. 5.**
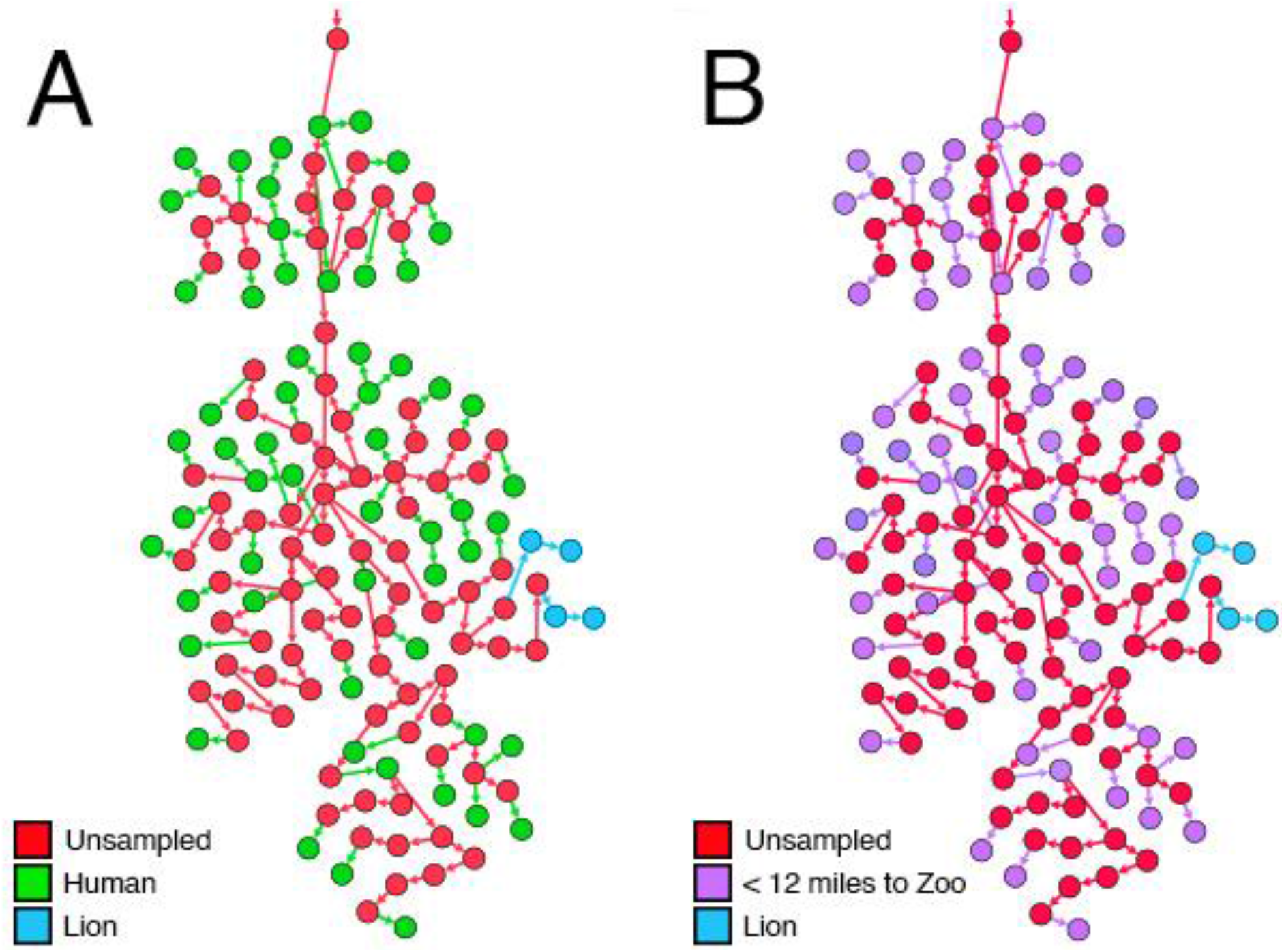
The prediction of the cross-species human to lion transmission of the SARS-CoV-2 virus. (A) Colour coded based on the host organism. It can be observed that the virus has been transmitted from a cluster of human hosts (green) to the lions (blue). Two lions have been initially infected from the human hosts and they, in turn, have infected the other lions. (B) Colour coded based on the vicinity of the sampled region from the Bronx Zoo, New York City. The entire cluster in that region is from areas that are less than 12 miles from the zoo which is typically less than a half-hour drive.

## 4. Discussion

Our approach combined Bayesian phylogenetics with TransPhylo’s ability to infer transmission networks complete with unsampled sources of infection to create a new and more robust pipeline than what is conventionally available. Such a pipeline can enhance current contact tracing approaches.

Through the repeated iterations and validation described above, we settled upon the five-step phylogenetic approach. The parameter optimised pipeline successfully predicted key transmission events using solely genomic data. The main inconsistencies in the pipeline were observed in BEAST2 prediction of “become uninfectious rate” and “reproductive number”. It should be noted that this inconsistency could be caused by the comparatively small sample size present as well the limited sample collection time available. Traditionally data spanning many years is used for such work, but this was unavailable in the COVID pandemic. We believe that these discrepancies can be resolved by increasing the sample size and collection time. Regardless of these challenges, the final output being the inference of transmission networks in a population appeared unaffected and consistent with the literature.

The ability to infer the presence of unsampled sources of infection leads to a number of exciting potential advantages when tackling epidemic control. Through the optimization of TransPhylo for COVID19, we have been able to explore this potential as can be seen in the New York State dataset. TransPhylo is capable of working on the assumptions that the pandemic is currently ongoing and future transmissions may continue to occur even after the completion of sample collection. It also assumes that the provided timed phylogenetic tree is accurate. Therefore, using a Bayesian approach, we have taken great lengths to ensure that our pipeline produces the most statistically sound tree. Based on these assumptions and the optimization of the tools’ sampling and infectious rates according to the region of study, we were able to infer complete transmission networks.

In all of our tests, the pipeline was able to successfully infer the transmission of the SARS-CoV-2 virus and gave a clear perspective of the pandemic’s progression. The consistency of the pipelines’ claims with that of the available literature proves that its predictions are reliable. In the analysis of the Russian and Canadian datasets, we observed the pipeline accurately predicts the progression of the pandemic. We were able to clearly see the countermeasures applied by relevant health care authorities take effect in our transmission diagrams.

The advantages of the power of predicting unsampled sources of infection to complete a transmission network becomes truly evident when handling problematic data such as the New York State data set. At the time of data collection, New York State was a COVID19 epicenter where the rate of infection had surpassed the sampling rate. The population was highly mobile with foreigners entering the state as well as locals traveling to and fro [39,41]. To further test the robustness of the pipeline we had introduced the sampled SARS-CoV-2 sequences obtained from the lions at the Bronx Zoo. Remarkably, the resultant transmission network diagram was able to clearly demarcate the mobile population and connect the scattered nodes through the inference of unsampled sources of infection. The pipeline was able to use partial genomic and epidemiological data to paint a complete picture of the pandemic’s status in the state. This was taken a step further when it was able to accurately infer the inter-species transmission of the virus from humans to lions. The validity of the result was further cemented since the surrounding nodes of sampled sources were from the neighbouring areas surrounding the Bronx Zoo. This cross-species transmission had plagued health care workers till it was validated by extensive laboratory testing [15]. But the pipeline was able to predict this transmission event solely from the available genomic data.

It needs to be noted that with unsampled individuals in our analysis, we mean that no sequence data has been obtained. These individuals could still have been tested and have been found positive for SARS-CoV-2. Combining the unsampled numbers inferred by our pipeline with the number of SARS-CoV-2 positive test numbers in the population would give insight into the number of undiagnosed and potential asymptomatic individuals in the population.

The purpose of the proposed pipeline is to reconstruct the transmission patterns in a population and establish a clear comprehensive picture of an infected population despite incomplete sampling. The pipeline’s purpose is to allow, using existing testing data for the optimisation of resources for epidemic control. We believe that this robust pipeline can truly be a front-line tool in the battle against infectious diseases.

## 5. Conclusion

In conclusion, we propose a new pipeline capable of computational inference of COVID19 transmission dynamics in a population complete with the direction of disease transmission and the prediction of unsampled sources. The development and execution of this pipeline can help to explain and confirm the expansion of the COVID19 pandemic. Through consequent refinement, the pipeline can be verified, improved, and ideally be used as a tool in the battle against COVID19. Due to the flexible nature of the pipelines guiding parameters, it holds the ability to be extended to other viral diseases.

## Data Availability

All sequence data is openly and readily available on GISAID. All software used are open-source.

https://github.com/theLongLab/TransCOVID

## All Codes for the Analyses can be Found Under

Project name: TransCOVID, Project home page: https://github.com/theLongLab/TransCOVID, Operating system(s): Platform independent, Programming language: R version 3+, License: MIT License, no restrictions for usage.

